# Identifying Online Advice-Seekers for Recovering from Opioid Use Disorder

**DOI:** 10.1101/2021.08.25.21262427

**Authors:** Gian-Gabriel P. Garcia, Ramin Dehghanpoor, Erin J. Stringfellow, Marichi Gupta, Jillian Rochelle, Elizabeth Mason, Toyya A. Pujol, Mohammad S. Jalali

## Abstract

**Background:** Online communities can provide social support for those recovering from opioid use disorder. However, advice-seekers on these platforms risk exposure to uncurated medical advice, potentially harming their health or recovery efforts. The objective of this analysis is to combine text annotation, social network analysis, and statistical modeling to identify advice-seekers on online social media for buprenorphine-naloxone use and study their characteristics.

**Methods:** We collected 5,258 posts and their comments from Reddit between 2014 and 2019. Among 202 posts which met our inclusion criteria, we annotated each post to determine which were advice-seeking (n=137) and not advice-seeking (n=65). We also annotated each posting user’s medication use stage and quantified their connectedness using social network analysis. In order to analyze the relationship between advice-seeking with a user’s social connectivity and medication use stage, we constructed four models which varied in explanatory variables.

**Results:** The stepwise model (containing “total degree” (P=0.002), “using: inducting/tapering” (P<0.001), and “using: other” (P=0.01) as significant explanatory variables) outperformed all other models. We found that users with fewer connections and who are currently using buprenorphine-naloxone are more likely to seek advice than users who are well-connected and no longer using the medication, respectively. Importantly, advice-seeking behavior is most accurately predicted using a combination of network characteristics and medication use status, rather than either factor alone.

**Conclusions:** Our findings provide insights for the clinical care of people recovering from opioid use disorder and the nature of online medical advice-seeking overall. Clinicians should be especially attentive (e.g., through frequent follow-up) to patients who are inducting or tapering buprenorphine-naloxone or signal limited social support.

## BACKGROUND

Many people recovering from opioid use disorder (OUD) face institutional barriers to recovery, including insufficient access to OUD treatment due to lack of health insurance and/or resources (e.g., transportation or time). For several reasons (e.g., poor treatment by healthcare professionals in prior care-seeking experiences), mistrust of medical professionals is common among those recovering from OUD [1, 2]. In addition, the stigma around OUD may cause providers to discriminate against those seeking medical treatment, potentially increasing mistrust between medical professionals and people who use opioids (PWUO) [3, 4]. Thus, even those who seek treatment may turn to online communities and social media platforms for the treatment advice and social support that they don’t find from their medical professionals.

Social media platforms such as Twitter, Facebook, and Reddit have fostered communities that provide solidarity and support for people dealing with a multitude of issues such as eating disorder recovery [5], suicidal ideation [6], chronic illnesses, and OUD [7]. In these communities, users can receive emotional support, information, and companionship while, in some cases, preserving anonymity. For PWUO, in particular, the anonymity of online forums, such as Reddit, has the potential to reduce stigma and social exclusion and can be an important factor for seeking support online [8, 9]. While these communities can provide much-needed support for those recovering from OUD, there is also an abundance of medical advice from non-clinicians (e.g., unverified OUD treatment alternatives [10]). Given the potential benefits and risks of seeking support for OUD online, it is important to identify and characterize which people recovering from OUD are likely to seek advice online. Identifying common characteristics among advice-seekers on these online platforms can also aid clinicians in proactively anticipating and addressing the needs of such PWUO.

Despite the importance of characterizing advice-seekers on online recovery platforms, there is currently limited research on this topic. Research has analyzed these platforms to predict PWUO’s transition to OUD [11], discover alternative treatments for opioid use recovery [10], and determine the prevalence of polydrug use [12]. Other work focuses on the social aspect of OUD recovery such as the social connectedness of online communities. For instance, similar to in-person support groups such as Alcoholics Anonymous and Narcotics Anonymous (AA/NA) [13], community engagement and cohesiveness in an online addiction recovery group is driven by its core of long-standing members [14]. Similarly, in another study using an online health forum for OUD recovery, people on one online health forum for OUD recovery were most engaged with the platform when they were “withdrawing” or “using” [15]. Despite the growing research in this space, little investigation has been done to better understand the connection between how a user’s advice-seeking behavior on an online platform is related to their online social connectedness and OUD buprenorphine-naloxone use stage.

To address this research gap, we identify the characteristics of PWUO who use online platforms for seeking medical advice for OUD recovery. Specifically, we analyze user and social network attributes of a community on Reddit with a focus on discussions related to Suboxone® (i.e., a brand name for buprenorphine-naloxone, and the most commonly discussed brand on Reddit), an effective medication used to support remission from, and prevent relapse to, OUD [16]. We combine text annotation, social network analysis, and statistical analysis to quantify the relationship between advice-seeking, buprenorphine-naloxone use stage, and social connectedness within this niche online community. Our study intends to improve the understanding of those who are most likely to seek OUD buprenorphine-related advice from online platforms.

## METHODS

### Data Description

Our data consisted of posts and comments collected from the “r/suboxone” subreddit, i.e., sub-community of Reddit described as “a community for all things buprenorphine.” We collected data from r/suboxone spanning February 4, 2014 (the inception of this subreddit) to December 31, 2019, excluding content created after January 1, 2020, to mitigate the potential effects of the COVID-19 pandemic on Reddit users’ posting behavior. We used the pushshift.io API [17] to collect URLs from all posts in this time period and used RedditExtractoR [18] library in R to extract relevant data and subsequent comments from each post.

### Exclusion Criteria and Data Sampling

To extract the most relevant posts for our analysis, we first excluded all empty and deleted posts since no text can be extracted from them, and network characteristics cannot be computed for users with deleted accounts. We then excluded all posts made by authors without one prior post, since users with no prior post-activity would have no connections to other users, resulting in an empty social network. Finally, because we were interested in medical advice-seeking, we narrowed our study sample to posts mentioning specific doctor/provider-related or Suboxone-related keywords, and the comments associated with these posts.

### Annotating Advice-seeking Posts and Suboxone Use Stage

With the final study sample, each post was annotated as advice-seeking or non-advice-seeking, in addition to the posting user’s Suboxone use stage. Three bachelor’s level research assistants [MG, EM, JR] and one Ph.D. student [RD] with backgrounds in medical informatics (all supervised by a substance use services researcher with expertise in qualitative coding and analysis [ES]) each annotated an initial 10 posts and collaboratively defined the criteria for the advice-seeking and Suboxone use stage criteria. A post was designated as “advice-seeking” if the user asked a specific question in their post about addiction, Suboxone, or doctor-related issues. For annotating the Suboxone use stage, three categories were considered: *using Suboxone, used to be on Suboxone*, and *cannot discern*. A user was annotated as “using Suboxone” if the content of their post indicated they were actively using Suboxone. Users who were identified as “using Suboxone” were further classified as “inducting,” “tapering,” or “other.” The inducting and tapering stages are critical points in the medication for opioid use disorder (MOUD) treatment process [19]. For people who use Suboxone, the inducting and tapering stages are well-known to be critical points in the MOUD treatment process [19]. A user was annotated as “inducting” if they had just begun or were about to begin Suboxone treatment, “tapering” if they were decreasing their dosage of Suboxone with the intention to stop taking Suboxone, and “other” if they were neither inducting nor tapering. For simplicity, we combined inducting and tapering into a single “inducting or tapering” category since both categories comprise transition stages. Users were annotated as “used to be on Suboxone” if they mentioned past use of Suboxone but have since stopped the treatment, or “cannot discern” if they did not give enough details to discern their Suboxone use stage. Uncertainties regarding annotations for specific posts were discussed and deliberated. All annotations were completed in Microsoft Excel.

### Measuring Social Connectedness

To characterize social connectedness, we constructed a social network graph for each sampled post based on a timeframe defined by the posting user’s first post or comment on r/suboxone and ended on the date at which they made the sampled post. All posts and comments made outside of this time period were not considered.

To construct each post-defined social network graph, we modeled nodes as unique users and edges as relations between two users. We added a directed edge, i.e., a relation, from user B to user A if either: (1) user A created a post and user B commented on that post, or (2) user A commented on a post and user B replied to that comment. The weight of each edge is equal to the number of relations between the two users on r/suboxone.

For each posting user, we computed their life span, total degree, eigencentrality, closeness, authority score, and hub score based on the user’s network at the time they made their post. A user’s lifespan is the total number of days between their first post or comment on r/suboxone and the date at which they authored the sampled post. A user’s total degree is the total number of relations to and from that user. Eigencentrality measures how influential a node is within the network [20]. For example, a user who is connected to many “important” users (i.e., other users with high eigencentrality) will have a relatively high eigencentrality. Closeness is equal to the inverse of the average length of the shortest paths to/from all the other vertices in the graph [21]. In other words, a user who is “close” to all other users in the social network (e.g., through direct connections with all other users or having direct connections with users who have many direct connections to all other users) would have a high closeness score. Finally, a user’s authority score and hub score represent two related centrality measures [22]. In this context, users with high authority scores will tend to receive comments from other users who frequently reply to others’ posts. Likewise, the users who tend to reply to others’ posts will have high hub scores.

### Statistical Analysis

We computed the total number of posts and the mean and standard deviation (SD) for all network characteristics and the number and proportions of posts by Suboxone use stage. We then divided our data into advice-seeking and not advice-seeking posts and repeated this analysis. Differences between advice-seeking posts and not advice-seeking posts were analyzed using the Mann-Whitney U test for all numerical study variables and the Pearson’s Chi-squared test for the Suboxone use stage variables. To determine which specific Suboxone use stage categories were driving significant differences across the entire group, we also conducted a post-hoc Chi-squared analysis on expected residuals [23] using the Benjamini-Hochberg p-value correction for multiple comparisons [24].

To quantify the relationship between advice-seeking (vs. not advice-seeking) with a user’s social connectivity and Suboxone use stage, we constructed four Generalized Linear Models (GLMs) with logit link functions. In each model, the dependent variable is given by a binary variable representing whether a post is advice-seeking or not. The independent variables include the posting user’s network characteristics and Suboxone use stage, the latter being re-coded as a series of binary variables using one-hot encoding with “used to be on Suboxone” as the reference category.

The *stepwise model* aimed to identify a parsimonious set of independent variables through statistical variable selection (i.e., using forward-backward selection). We compared the stepwise model to three additional models: the full model, Suboxone use model, and network model. The *full model* contained all network characteristics and the Suboxone use stage variables. The Suboxone *use model* and *network model* contained only the Suboxone use stage and network characteristics variables, respectively. To aid our inference of modeling coefficients, we computed the variable inflation factor to assess the multicollinearity of modeling variables for each model. We then applied leave-one-out cross-validation to evaluate each model using Area Under the Receiving Operator Characteristic Curve (AUROC), Akaike Information Criterion (AIC), and F1 score. Altogether, these measures provide a holistic picture of each model’s predictive performance.

## RESULTS

### Data Characteristics

The final study sample contained 202 posts (Figure 1). Table 1 summarizes these data with respect to our study variables. Within these posts, 137 (67.8%) were advice-seeking.

**Figure 1:**
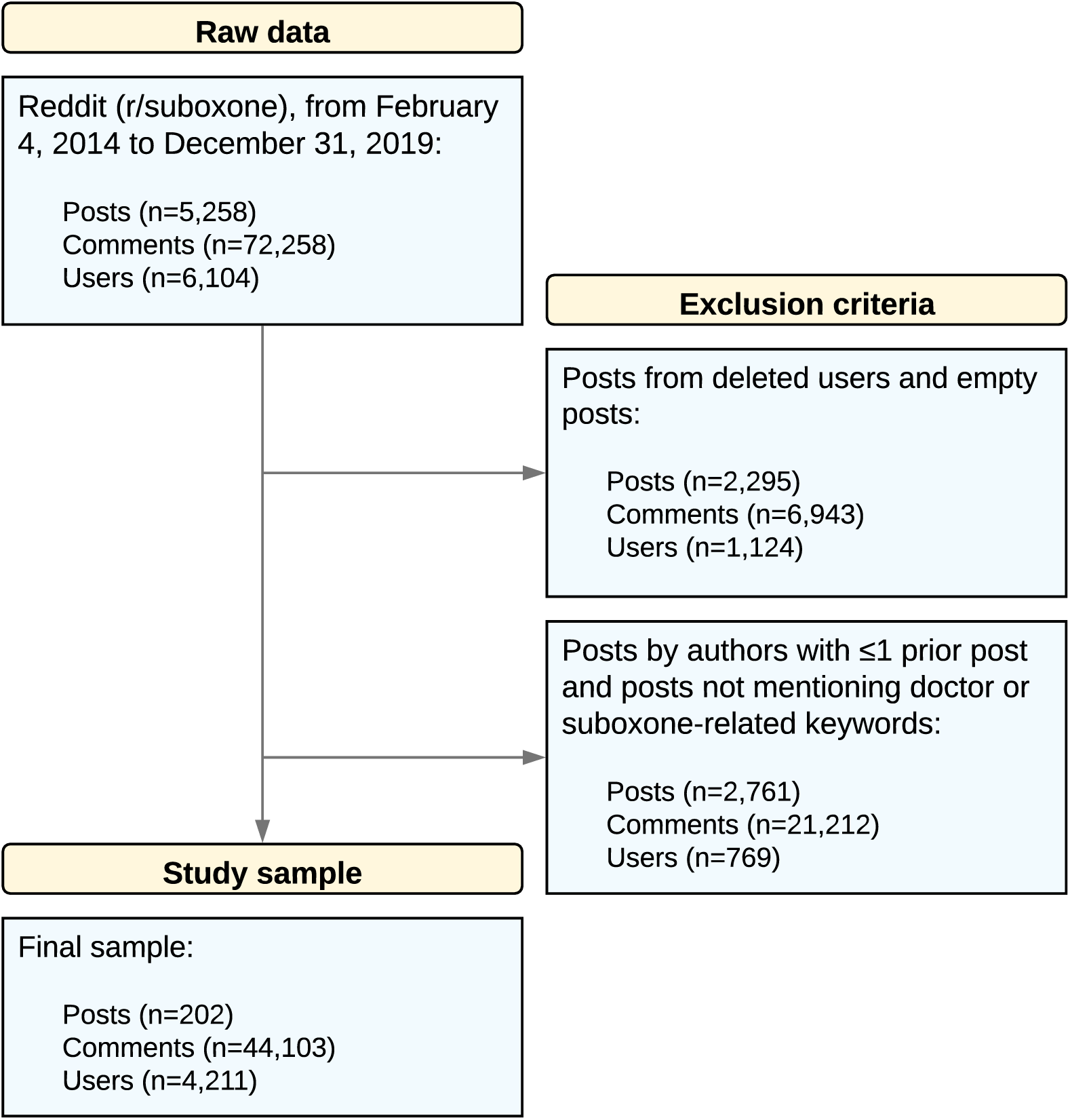
Application of exclusion criteria to raw data for obtaining the study sample

**Table 1:**
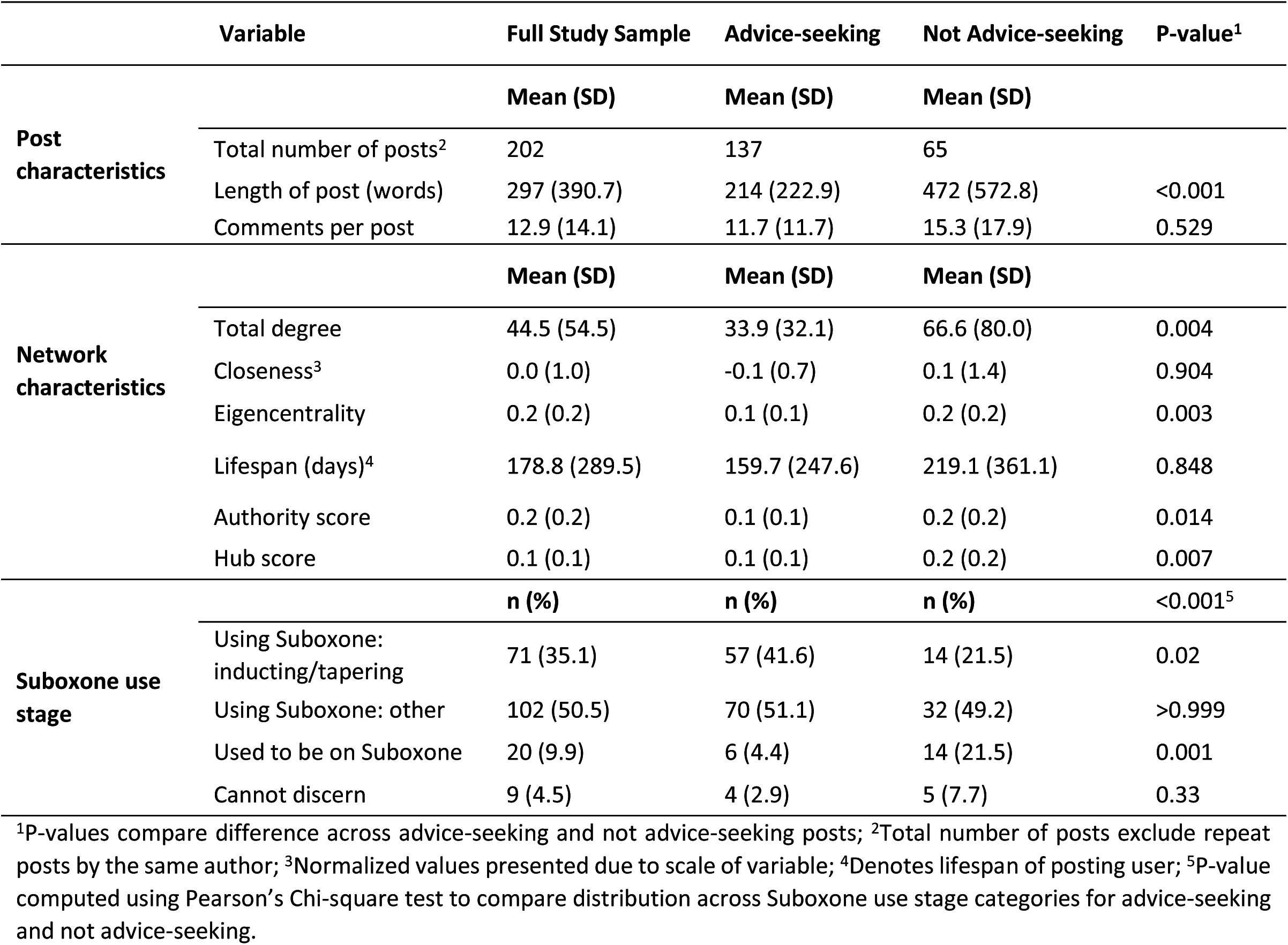
Description of data with respect to post characteristics, network characteristics, and Suboxone use stage

Among posting users, those who made advice-seeking posts had a significantly different total degree (P=0.004), eigencentrality (P=0.003), authority score (P=0.014), and hub score (P=0.007) than users who did not make advice-seeking posts. Additionally, the proportion of users in each Suboxone use stage was significantly different between users who authored advice-seeking posts and those who did not (P<0.001). Notably, there was a significantly greater proportion of advice-seeking users vs. not advice-seeking users who were inducting or tapering (n=57, 41.6% vs. n=14, 21.5%; P=0.02) and a significantly lesser proportion of users who used to be on Suboxone (n=6, 4.4% vs. n=14, 21.5%; P=0.001).

Figure 2 illustrates examples of advice-seeking and not advice-seeking posts, along with the posting user’s social network graph, post characteristics, network characteristics, and Suboxone use stage. These examples were selected to be close to the mean total degree in each category.

**Figure 2:**
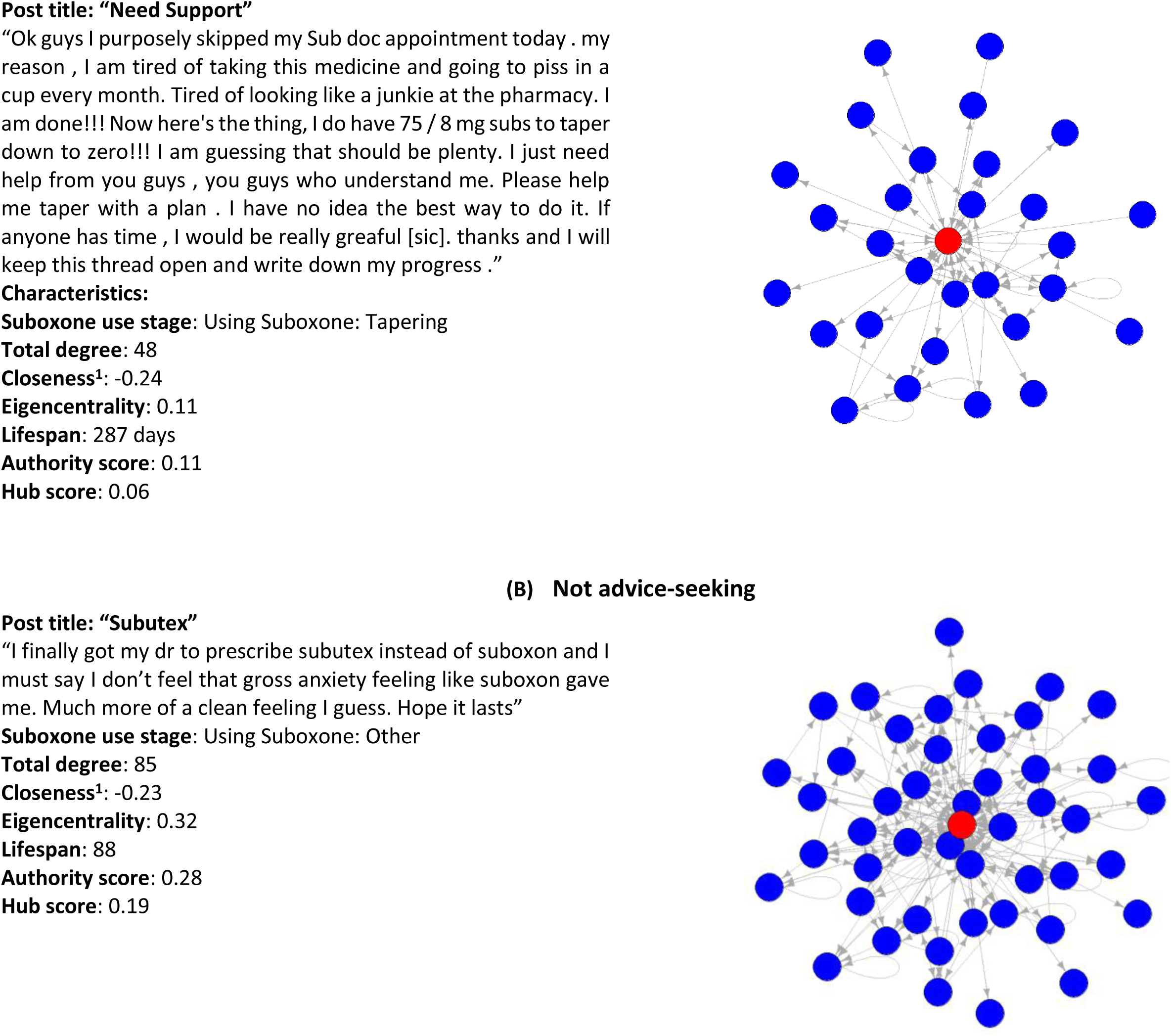
Illustration of (A) advice-seeking and (B) not advice-seeking posts with posting user’s network

Posting user shown as a red node, all other users are shown as blue nodes, edges indicate the relation between users. ^1^Normalized value computed by subtracting the sample mean and dividing by the sample standard deviation

### Regression Modeling

Our GLMs are described in Table 2. In the stepwise model, “total degree,” “closeness,” and the Suboxone use stage variables were selected by the variable selection procedure, with “total degree” (P=0.002), “using Suboxone: inducting/tapering” (P<0.001), and “using Suboxone: other” (P=0.002) being significantly different from 0 (i.e., strongly associated with advice-seeking). These three variables had variance inflation factors (VIF) ranging from 1.04-1.08, indicating low multicollinearity, and none of the variables that were removed by the stepwise variable selection procedure were significant in any of the other GLMs. Additionally, all variables that were significant in the stepwise model were significant in at least one other GLM. Notably, VIFs in all other models were low-moderate (i.e., VIF≤5) except for eigencentrality (VIF=10.94-11.26) and authority score (VIF=9.53-9.69) within the full and network models. Nevertheless, these variables had coefficient estimates close to 0 and were not significant in either model.

**Table 2:** Model coefficients and performance measures for each model predicting the likelihood of advice-seeking

| Model | Full | Network | Suboxone Use | Stepwise |
| --- | --- | --- | --- | --- |
| *Coefficient<sup>1</sup> (95% CI)* |  |  |  |  |
| Intercept | -0.86 (-2.07, 0.24) | 0.67* (0.08, 1.25) | -0.85 (-1.89, 0.07) | -0.69 (-1.76, 0.28) |
| Total degree | -0.82 (-1.70, -0.01) | -0.81* (-1.65, -0.06) |  | -0.63** (-1.07, -0.25) |
| Closeness | -0.33 (-0.83, 0.04) | -0.30 (-0.74, 0.05) |  | -0.31 (-0.69, -0.01) |
| Eigencentrality | 0.00<sup>†</sup> (-1.18, 1.20) | -0.02<sup>†</sup> (-1.20, 1.12) |  |  |
| Lifespan | 0.00 (0.00, 0.00) | 0.00 (0.00, 0.00) |  |  |
| Authority score | -0.05<sup>†</sup> (-1.15, 1.07) | -0.02<sup>†</sup> (-1.13, 1.09) |  |  |
| Hub score | 1.69 (-2.77, 6.39) | 1.18 (-3.19, 5.80) |  |  |
| Using Suboxone: inducing/tapering<sup>2</sup> | 2.10*** (0.96, 3.34) |  | 2.25*** (1.17, 3.44) | 2.08*** (0.94, 3.31) |
| Using Suboxone: other<sup>2</sup> | 1.41* (0.33, 2.59) |  | 1.63** (0.63, 2.74) | 1.42* (0.35, 2.57) |
| Cannot discern<sup>2</sup> | 1.02 (-0.83, 2.92) |  | 0.62 (-1.04, 2.28) | 1.09 (-0.67, 2.90) |
| **Performance measures<sup>3</sup>** |  |  |  |  |
| AUROC | 0.61 | 0.54 | 0.52 | 0.66† |
| AIC | 239.19 | 246.85 | 241.02 | 231.86† |
| F1 | 0.44 | 0.30 | 0.40 | 0.47† |

<sup>1</sup>significance: \*<0.05 \*\*P<0.01, \*\*\*P<0.001; <sup>2</sup>Suboxone use stage variable binarized using one-hot encoding with “Used to be on Suboxone” as the reference category; <sup>3</sup>AUROC: Area under the receiver operating characteristic curve; AIC: Akaike Information Criterion; †best-performing model; †variance inflation factor>5.

Whether each variable increased/decreased the likelihood of being an advice-seeker (i.e., whether the coefficient was positive/negative) was consistent across all models. Among variables with coefficients significantly different from 0, “total degree” had a negative coefficient, indicating that posting users with more connections were less likely to be advice-seeking. Likewise, the coefficients for “using Suboxone: inducting/tapering” and “using Suboxone: other” were positive, indicating that posting users who were identified as using Suboxone were more likely to be advice-seeking than users who were identified as formerly using Suboxone.

With regard to performance measures, the stepwise model outperformed all other models with the greatest AUROC (0.66 vs. 0.52-0.61), least AIC (231.86 vs. 239.19-246.85), and greatest F1 score (0.47 vs. 0.30-0.44).

## DISCUSSION

In this research, we assessed the relationship between advice-seeking behavior, social connectedness, and buprenorphine-naloxone (Suboxone®) use stage in Reddit. While previous studies using Reddit data have individually analyzed social roles and connectedness of subreddits [9], advice seeking among users with OUD [25], and posts regarding induction and tapering [26], our study is the first to connect the three topics and to do so by combining social network analysis, text annotation, and statistical modeling.

Our analysis focused on advice-seekers in r/suboxone. Suboxone (and buprenorphine and/or buprenorphine-naloxone, more broadly) is an effective treatment for OUD [16, 19, 27]. Nevertheless, there remain many limitations to Suboxone treatment (e.g., insufficient training of medical professionals and stigma) that erode trust between PWUO and their providers [28, 29]. These limitations may lead PWUO to seek treatment advice online, as evidenced by the subreddit we analyzed. Our analysis provides some insight into the characteristics of advice-seekers on Reddit, potentially identifying PWUO who may not be receiving sufficient medical advice or support from their prescribing clinician.

In our best-performing model (i.e., stepwise), we found that advice-seeking behavior had significant associations with a user’s total degree (i.e., number of connections with other users) and Suboxone use stage. In particular, our results suggest that users with few connections on r/suboxone as well as those who are using Suboxone are more likely to seek advice online than users who have more connections and no longer using Suboxone. Regarding advice-seeking among Suboxone users, our descriptive analysis proved a significantly higher proportion of advice-seekers who were inducting or tapering compared to non-advice-seekers. This result may suggest that PWUO who are inducting or tapering, i.e., in a transition stage, may be more likely to seek advice online than Suboxone users who are not in a transition stage. To this end, previous research has connected social network characteristics with advice-seeking in addiction recovery homes and found that the number of advice-seeking relationships was positively correlated with high levels of stress [30]. While we did not explicitly study stress, previous literature has reported increased stress levels among patients who are initiating or tapering with buprenorphine, partly due to intense withdrawal symptoms [31, 32]. Thus, stress may play a key role in the positive relationship between being in a transition stage and advice-seeking—future research should investigate this assumption especially in social media platforms. In stressful circumstances, people who are seeking advice might feel more comfortable doing so from their peers, rather than their providers, especially if they remain ambivalent about quitting and are concerned about judgment from their providers [33, 34].

While we found no link between lifespan (i.e., the length of time a user has been active on r/suboxone) and advice-seeking behavior, we did find a relationship between Suboxone use stage and advice-seeking behavior. In this way, our results suggest similar dynamics to those in AA/NA, where new members are encouraged to seek advice and support from more experienced sponsors [13]. In the context of an anonymous online forum, inducting onto Suboxone (a subset of our analyzed group) is a better corollary to the AA/NA “newcomer” than lifespan per se.

From a clinical perspective, our findings suggest the need for prescribing clinicians to pay special attention (e.g., by providing more frequent follow-up and being especially accessible) to PWUO who are inducting or tapering and who have a limited support network. The importance of having a strong support network for maximizing the likelihood of recovery is well-established [19, 35]. However, how to anticipate and address the potential needs of PWUO in the inducting or tapering stage remains challenging for clinicians; mention of these transition stages is typically restricted to recommendations to help clinicians determine dosing levels or identify when it is appropriate to begin inducting or tapering with buprenorphine [19, 36, 37]. As clinical best practices and public health interventions for OUD treatment continue to evolve, it will be critical to understand why PWUO in transition stages turn to online platforms, what specific advice they are seeking, and whether some or all of their needs could be better addressed by providers.

Beyond the clinic, quickly and accurately identifying advice-seeking users can help online platforms automate the delivery of medically sound informational resources (e.g., via chatbots [38]) for people recovering from OUD. Since our stepwise model achieved greater predictive accuracy than the network and Suboxone use models, these results suggest that the combination of network characteristics and Suboxone use stage are better indicators of advice-seeking behavior than either of those factors individually. Notably, the network model attempted to include a more comprehensive description of each user’s network compared to the stepwise model. However, none of the additional variables beyond total degree were significant and the network model had far worse predictive performance than the stepwise model. These findings indicate the importance of focusing on the right measures of social connectedness when attempting to identify advice-seekers on online platforms. Fortunately, the total degree is relatively simple to compute. Hence, if Suboxone use stage can be classified with relatively high accuracy (e.g., using natural language processing methods), then our stepwise model can provide a starting place for identifying users who might benefit from targeted medically-sound advice on online platforms. Research into whether users of online platforms would welcome such advice is warranted.

Our findings may also be connected with the use of online platforms for other health areas. Online information seeking, especially for chronic diseases, weight loss, and mental health issues, is commonplace [5–7]. Across many health areas and online platforms, drivers for seeking health information online include gaining social support [39, 40] and receiving tailored advice from online users with similar experiences [41, 42]. While an analysis of the contents of our post data was beyond the scope of this research, it is plausible that advice-seekers on r/suboxone were also hoping to gain social support or receive individualized treatment advice.

This research is not without its limitations. First, this study focuses only on advice-seeking on r/suboxone. Future research can consider additional social roles, including users who give advice or social support, on additional opioid-related subreddits such as r/opiates. Second, our study focused on the characteristics of advice-seeking users and not the characteristics of the posts themselves.

Additional insights can be drawn from analyzing the content of the posts to highlight patterns of advice-seeking posts and facilitate the automated identification of advice-seekers. This analysis could even be extended to evaluate the quality of advice shared on these online platforms. Third, our work leveraged manual annotations of users’ posts to determine respective Suboxone use stages. Future research may explore algorithmic techniques to classify such users, which in tandem with the model presented here would streamline the identification of advice-seeking users and facilitate analysis of topics for which advice is often sought. Finally, our study is limited to data ending on December 31, 2019. The onset of COVID-19 has brought many challenges to PWUO, which could have changed the nature of their online activity and interactions. Thus, future analyses can investigate how online social roles may have changed since the pandemic started in the United States.

Despite these limitations, this research: (1) demonstrates a method to classify advice-seeking users based on their network characteristics and buprenorphine-naloxone use stage; (2) sheds light on the characteristics of advice-seekers on an online platform for OUD recovery; and (3) provides insights for the clinical management of PWUO who are recovering from OUD as well as the nature of online medical advice-seeking. Given the vulnerability of PWUO, it is imperative that future research continues to explore the needs of this population and how they can be met.

## Data Availability

Data can be requested from the corresponding author.

